# Long-Term Outcomes of Untreated Micropenis: Growth Patterns and Predictive Factors

**DOI:** 10.1101/2024.07.21.24310773

**Authors:** Davoud Amirkashani, Mostafa Abdollahi, Mostafa Masoumi

## Abstract

**Background:** Micropenis, defined as a penile length more than 2.5 standard deviations below the mean for age and population, presents significant concerns for patients and parents. Despite current guidelines recommending multidisciplinary management, there is limited evidence on long-term outcomes, particularly in untreated patients.

**Methods:** This retrospective cohort study involved 46 male children aged 7 to 9 years presenting with micropenis at the Ali Asghar Endocrine Clinic from 2015 to 2023. Initial penile size, BMI, and other growth parameters were measured, with biannual follow-ups extending three years post-bone fusion to evaluate growth rates and influential factors.

**Results:** Initial mean stretched penile length (SPL) was 3.22 ± 0.21 cm. Significant increases in penile size were observed across all intervals, with the highest growth rates occurring between the first- and second-years post-fusion. BMI emerged as the most significant predictor of penile growth, while initial SPL was the least influential factor. By the third-year post-fusion, all subjects achieved penile lengths within the normal range.

**Conclusion:** Our findings indicate that most untreated micropenis patients attain normal penile size by adulthood, highlighting the importance of monitoring growth rates rather than focusing solely on initial penile size. This study provides critical insights for developing guidelines and management strategies for micropenis, emphasizing the necessity of continued follow-up to ensure optimal outcomes.

## Introduction

Penile length is a significant aspect of male anatomical and physiological health (such as sexual development, impacting not only sexual function but also psychological well-being and social dynamics or even regional cultures across time(1-3).

Previous studies have shown that factors such as BMI (Body Mass Index), age of puberty onset, and the timing of bone fusion can significantly influence penile development and length(4, 5). These findings highlight the multifaceted nature of factors affecting penile growth and development. Additionally, penile length can serve as an indicator of various malformations, including hormonal imbalances, congenital anomalies, metabolic conditions, and anatomical defects (6).

With all that, there is an uprising of concern among parents and patients about penile length (1, 7). As a result, urologists and endocrinologists are facing an increasing number of patients with concerns about short penis. The majority of these individuals have penile lengths that fall within the normal range according to established guidelines and do not require medical intervention (1). However, a subset of these patients lies in the micropenis rage and do present with genuine concerns that show the importance of further evaluation and potential treatment.

Micropenis is defined as a penile length more than 2.5 standard deviations below the mean for age and population (8). Current guidelines recommend a multidisciplinary approach involving endocrinologists, urologists, and psychologists, with treatments often such as hormonal therapy (6, 8, 9). However, there is a lack of robust evidence supporting the long-term efficacy and safety of these treatments and also the outcome of patients who didn’t receive the treatment. Additionally, existing guidelines do not provide clear criteria for when or how physicians should initiate or withhold treatment, underscoring the need for comprehensive guidelines that include long-term follow-up to provide valuable and consistent care for patients with micropenis, particularly those who have not undergone treatment (1).

As we recognize the gap in studies among patients not receiving treatment, our aim is to longitudinally follow up these patients from their initial assessment through post-puberty, examining the impact of somatometric factors. Additionally, we aim to demonstrate that patients who do not receive treatment often achieve normal penile size by the conclusion of the study period.

## Method

### Study design

We enrolled 90 male children, aged 7 to 9 years, presenting with micropenis as the chief complaint brought by their parent. This retrospective cohort study was conducted at the Ali Asghar Endocrine Clinic from 2015 to 2023 (visiting twice a year for follow up). None of the boys included had congenital anomalies, metabolic diseases or any other background visits (1).

All boys who met the criterion of micropenis by having a stretched penile length shorter than the 2.5 SD less than the mean length, according to “the 2011 age-matched New York Cohen children medical centre of north shore-long island”(10), penile length standards, were included in the study (2, 11, 12).

All measurements were conducted by the same observer (first author, NT). The prepubic subcutaneous fat at the base of the penis was compressed with one end of the ruler down to the pubic ramus. The observer then fully stretched the penis by holding the glans between the left thumb and index finger, while the ruler was placed along the side of the stretched penis using the observer’s right hand. The length from the lower edge of the pubic bone to the tip of the glans (excluding foreskin) was recorded. It’s important to mention that all of the subjects were circumcised.

Based on the 6 months visit schedule, in the first visit we documented the weight, height and calculated BMI and age at the first visit. During subsequent visits, testicular size and volume were assessed by the same observer (first author). The onset of puberty was determined when testicular size reached 2.5 cm or a volume of 4 cc, as measured using an orchidometer. Additionally, the age of bone fusion was determined by examining the growth charts (Iran National Growth Chart provided by the Ministry of Health of Iran) and using wrist x-ray imaging when the growth rate approached zero or plateaued. Patients were followed up for three years after identifying the age of bone fusion, with assessments conducted at one-year intervals, during which penile size was measured.

### Ethical considerations

At the beginning of the study, the research protocol was thoroughly explained to the parents, and informed consent was obtained. During each visit or examination, one or both parents were present to observe the examination.

The protocol for this study was reviewed and approved by the Institutional Review Board of Iran University of Medical Sciences (approval number: 05-2024-155).

### Statistical analysis

Quantitative variables are expressed as mean ± standard deviation, while categorical variables are presented as frequency (percentage). Statistical analyses were conducted using the t-test and linear mixture model with the Python statistical libraries NumPy 2.0.1 (13) and SciPy1.140. A p-value less than 0.05 was considered statistically significant.

## Results

### 1. Characteristics of patients

The characteristics of the patients presenting with concerns regarding micropenis are detailed in Table 1. A total of 46 patients, with an age range of 7 to 9 years (mean ± SD, 8.04 ± 0.77 years), were included in the study. The initial stretched penile length (SPL) for all patients was measured at 3.22 ± 0.21 cm. (Table-1)

**Table-1).** Patients Characteristics.

Figures/Tables
| Charasterics | Min | Max | Median | Mean | Standar Deviation (std) |
| --- | --- | --- | --- | --- | --- |
| First Visit Age (year) | 7 | 9 | 8 | 8.04 | 0.77 |
| Height (cm) | 113 | 135 | 124.5 | 125.02 | 6.07 |
| Weight (kg) | 18 | 34 | 24.5 | 25.34 | 3.92 |
| BMI | 12.71 | 20.43 | 16.19 | 16.15 | 1.67 |
| Size in First Visist | 2.8 | 3.8 | 3.2 | 3.22 | 0.21 |
| Puberty Age | 9 | 12 | 11 | 10.67 | 0.88 |
| Puberty Size | 4.2 | 6.4 | 5.4 | 5.45 | 0.44 |
| Fusion Age | 14.9 | 17 | 15.9 | 15.82 | 0.57 |
| Fusion Size | 6.9 | 14 | 11.05 | 11.14 | 1.33 |
| 1/y after Fusion | 7.5 | 14.6 | 11.6 | 11.76 | 1.24 |
| 2/y after Fusion | 7.5 | 15 | 13.65 | 13.31 | 1.37 |
| 3/y after Fusion | 7.9 | 15 | 14 | 13.91 | 1.32 |

As illustrated in Figure 1 and Table 2, there is a significant increase in penile size observed from the initial visit to the third-year post-fusion. Additionally, significant increments are evident in penile length from the first visit to puberty, continuing from puberty to fusion, and further progressing from fusion to the final follow-up visit. (Figure-1, Table-2)

**Figure-1).**
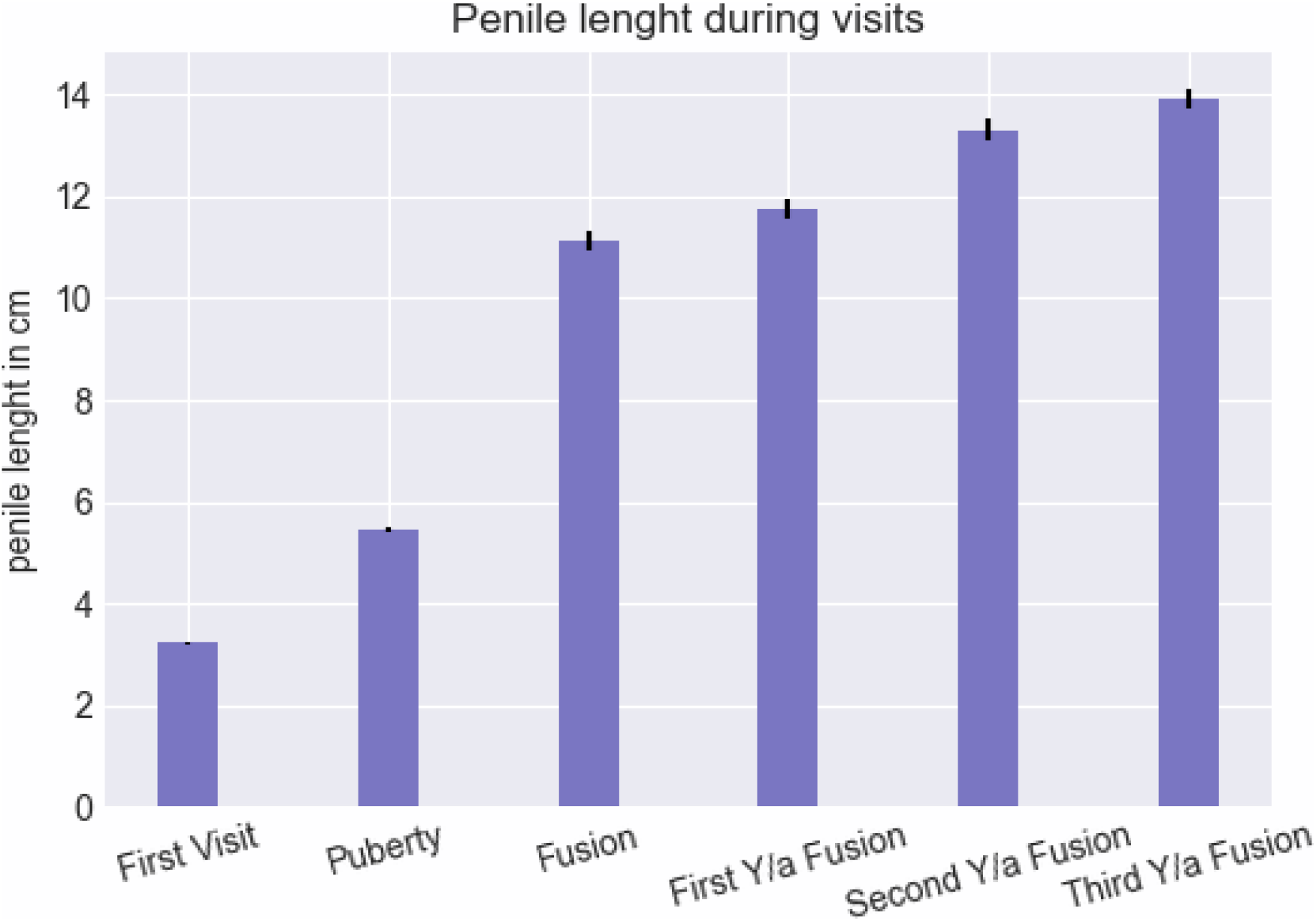
Comparing Penile length in different visits.

**Table-2).** Mean Penile lengths and Standard errors of visiting Checkpoints.

| checkpoints | mean penile length (cm) | Standard error |
| --- | --- | --- |
| First Visit | 3.22 | 0.03 |
| Puberty | 5.45 | 0.06 |
| Fusion | 11.14 | 0.19 |
| First Year/a Fusion | 11.76 | 0.18 |
| Second Year/a Fusion | 13.31 | 0.2 |
| Third Year/a Fusion | 13.91 | 0.19 |

As depicted in Figure 2 and Table 3, it is evident that all growth rates are statistically significant and differ significantly from each other. The most rapid increase in penile size occurs between the first and second years following fusion (1), while the smallest increment is observed between the second- and third-years post-fusion. (We dropped the size of penis as first, second and third year after fusion) (Figure-2, Table-3)

**Table-3).** Penile Growth rate per-year based on the checkpoints in cm.

| checkpoints | Growth rate per year (cm) |
| --- | --- |
| FV-Puberty | 1.02 |
| Puberty-Fusion | 1.17 |
| Fusion-1y/f | 0.62 |
| 1y/f - 2y/f | 1.54 |
| 2y/f -3y/f | 0.6 |

**Figure-2).**
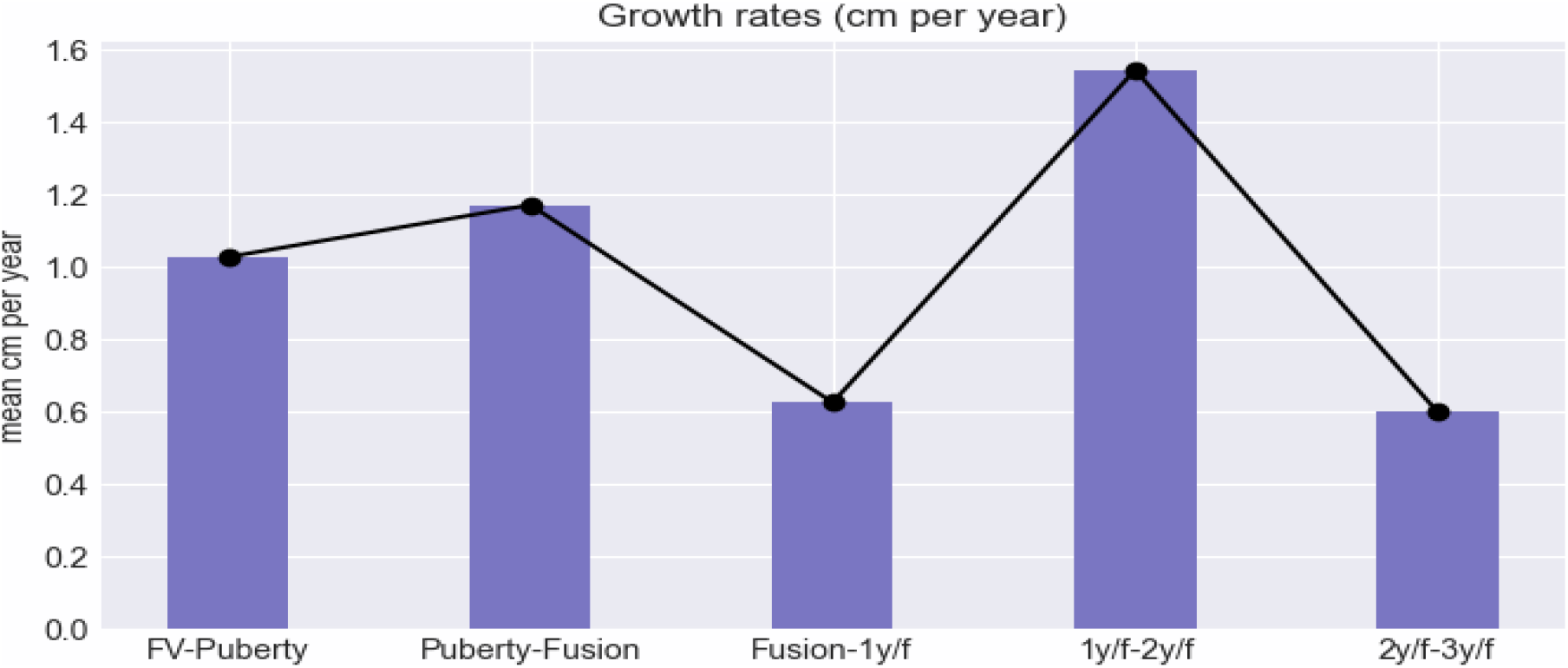
Penis Growth rates per year in cm

From Figure 3, the application of the linear mixture model facilitated the extraction of the most influential factors, with BMI emerging as the most significant predictor. Conversely, the initial penile size at the first visit was identified as the least influential factor in determining subsequent growth trajectories.

**Figure-3).**
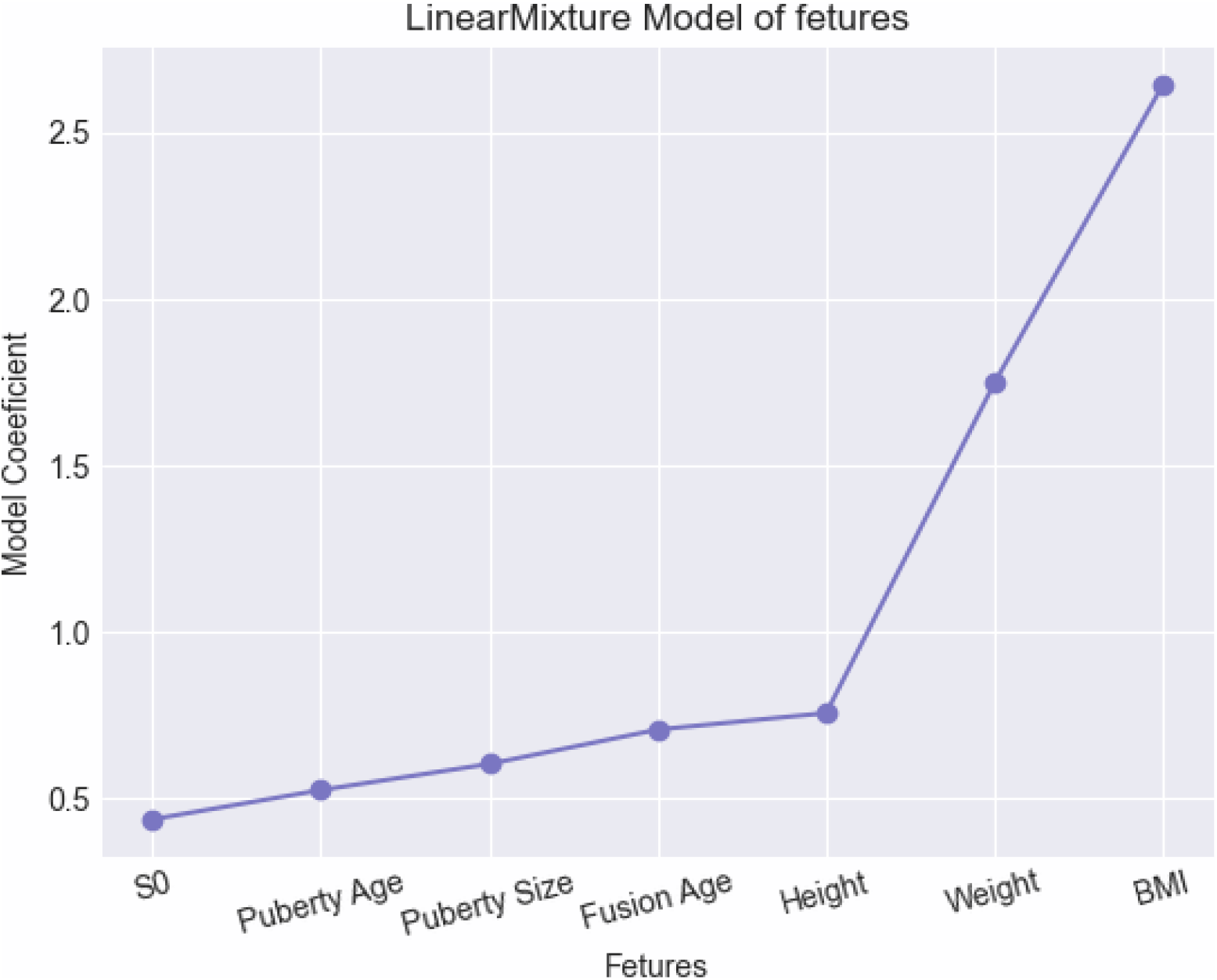
linear mixture model of important factors and their Coefficients

Figure 4 presents a correlation matrix that elucidates the relationships between various parameters. This heatmap allows for the visualisation and analysis of correlations among key variables, including BMI, age of puberty onset, hormonal profiles, and penile length measurements. The matrix provides valuable insights into the interdependencies and potential associations among these factors within the context of penile growth and development.

**Figure-4).**
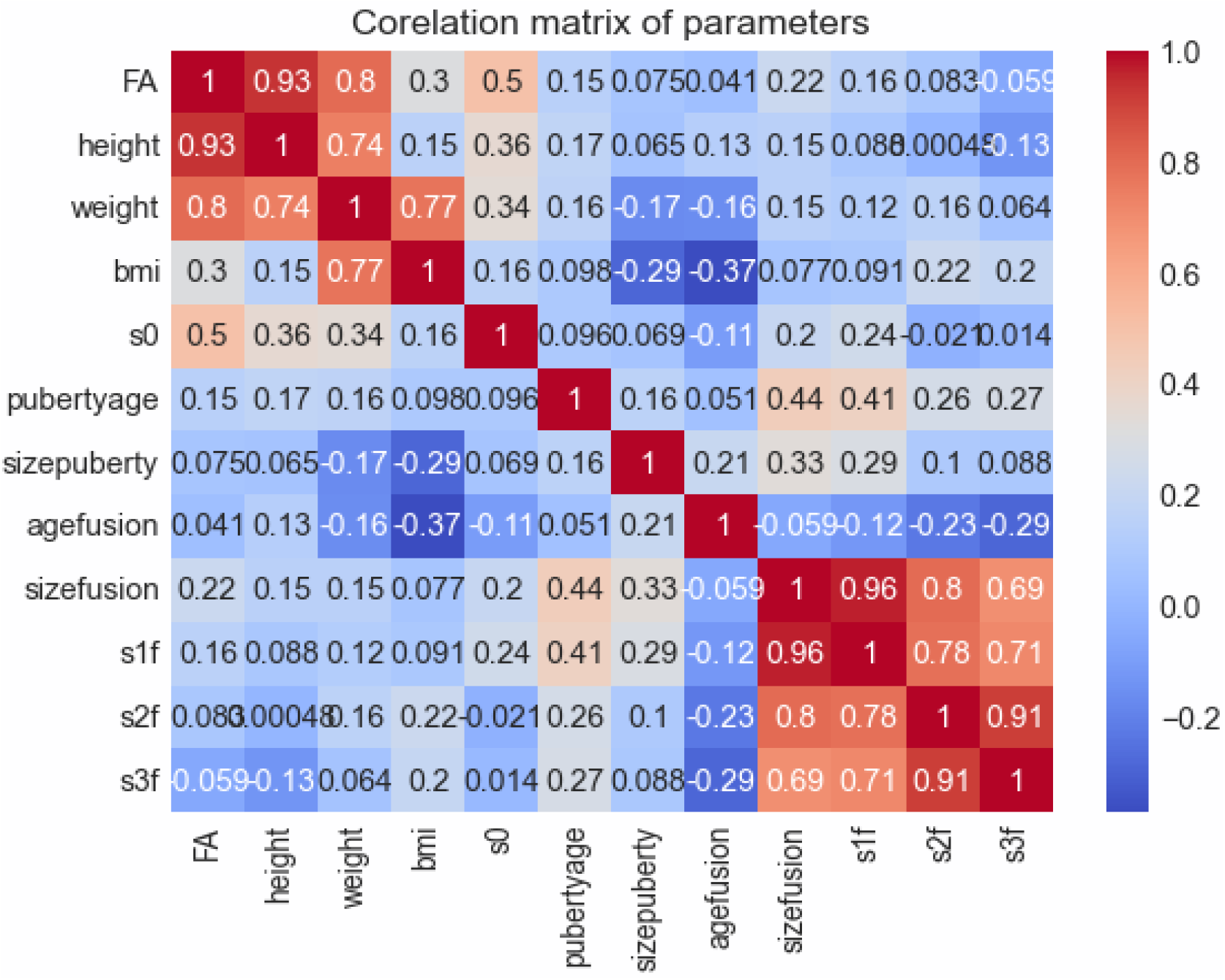
Correlation matrix of important variables

## Discussion

The aim of our study was to conduct a long-term follow-up of patients with micropenis who did not receive any treatment and to demonstrate that most of these patients will achieve normal penile size as they grow, including three years post-bone fusion. Our findings indicated that the entire population will reach normal penile size by puberty, as per the New York penile length chart (10). Furthermore, penile length continues to increase after puberty, extending into the post-bone fusion period (1). By the third-year post-fusion, our comparisons reveal that the entire cohort surpasses the minimal normal range of penile length based on Iranian penile length standards.

One of the most crucial factors in properly following up with patients is monitoring the growth rate of penile size during the intervals between visits. Our study identified that the highest growth rates occur between the first- and second-years post-fusion. The growth rates for other intervals, in descending order, are: puberty to fusion, the first visit to puberty, fusion to the first-year post-fusion, and lastly, from the second-year post-fusion to the third-year post-fusion. Although two subjects reached a normal penile size by puberty, their growth rates between puberty and bone fusion were insufficient compared to the population, underscoring the necessity of continued follow-up even after achieving normal penile size. This finding indicates to physicians that only a few patients with true micropenis will require treatment, emphasising the importance of ongoing monitoring to ensure proper development Additionally, physicians can use these growth rate intervals as checkpoints for patient follow-up.

As previously mentioned, there are correlation factors that assist physicians in predicting patient penile size growth and prognosis. These factors are crucial for determining the optimal timing for treatment initiation to achieve the best outcomes with minimal side effects during both childhood and adulthood. The most significant factor is BMI, which exhibits a strong correlation coefficient. The second most influential factor is the age of bone fusion. Conversely, the least effective factor among those measured is the penile size at the first visit.

There are several limitations in our study that should be mentioned. The first limitation is the study population. Due to the requirement for long-term follow-up and multiple visits, the number of participants decreased over the years (1). Secondly, there were exogenous factors beyond our control, such as nutritional and economic conditions, although no proven nutritional deficiencies were identified (1). Lastly, the study was conducted over seven years, preventing us from continuing follow-up for the fourth- and fifth-years post bone fusion.

Future studies should aim to conduct this research on a larger population (1) and consider a longer follow-up period, particularly extending into adulthood, to analyse long-term physical and psychological effects. Additionally, we recommend that future researchers establish comparative studies between our data and those of individuals receiving hormonal therapy.

This study offers valuable insights into penile growth among the micropenis population from the prepubertal to post-puberty stages. The growth rates and the correlation coefficients previously discussed provide a guideline and checkpoints for physicians, aiding in optimal patient management. The most significant finding of our study is that the growth rate of penile size, rather than the penile size itself, serves as the primary parameter of interest. This focus on growth rate is crucial for guiding clinical decisions and ensuring effective patient care.

## Data Availability

All data produced in the present study are available upon reasonable request to the authors.

## References

1. Han JH, Lee JP, Lee JS, Song SH, Kim KS. Fate of the micropenis and constitutional small penis: do they grow to normalcy in puberty? J Pediatr Urol. 2019;15(5):526.e1-.e6.

2. Kareem AJ, Elusiyan JBE, Kareem AO. Stretched penile length and total serum testosterone in term male neonates. Pan Afr Med J. 2020;37:61.

3. Soheilipour F, Rohani F, Dehkordi EH, Isa Tafreshi R, Mohagheghi P, Zaheriani SM, et al. The Nomogram of Penile Length and Circumference in Iranian Term and Preterm Neonates. Front Endocrinol (Lausanne). 2018;9:126.

4. Mancini M, Pecori Giraldi F, Andreassi A, Mantellassi G, Salvioni M, Berra CC, et al. Obesity Is Strongly Associated With Low Testosterone and Reduced Penis Growth During Development. J Clin Endocrinol Metab. 2021;106(11):3151–9.

5. Mushannen T, Cortez P, Stanford FC, Singhal V. Obesity and Hypogonadism-A Narrative Review Highlighting the Need for High-Quality Data in Adolescents. Children (Basel). 2019;6(5).

6. Becker D, Wain LM, Chong YH, Gosai SJ, Henderson NK, Milburn J, et al. Topical dihydrotestosterone to treat micropenis secondary to partial androgen insensitivity syndrome (PAIS) before, during, and after puberty - a case series. J Pediatr Endocrinol Metab. 2016;29(2):173–7.

7. Lee PA, Mazur T, Danish R, Amrhein J, Blizzard RM, Money J, et al. Micropenis. I. Criteria, etiologies and classification. Johns Hopkins Med J. 1980;146(4):156–63.

8. Hatipoğlu N, Kurtoğlu S. Micropenis: etiology, diagnosis and treatment approaches. J Clin Res Pediatr Endocrinol. 2013;5(4):217–23.

9. Boiko MI, Notsek MS, Boiko OM. The Efficacy of Injection Penile Girth Enhancement as an Option for Small Penis Syndrome Management. Aesthet Surg J. 2023;44(1):84–91.

10. Wiygul J, Palmer LS. Micropenis. ScientificWorldJournal. 2011;11:1462–9.

11. Teckchandani N, Bajpai M. Penile length nomogram for Asian Indian prepubertal boys. J Pediatr Urol. 2014;10(2):352–4.

12. Wood K, Nanduri V, Merchant N. Is it micropenis? Does size matter? Arch Dis Child Fetal Neonatal Ed. 2017;102(4):F345.

13. Harris CR, Millman KJ, van der Walt SJ, Gommers R, Virtanen P, Cournapeau D, et al. Array programming with NumPy. Nature. 2020;585(7825):357–62.

